# Cross-Sectional and Longitudinal Evaluation of Heart-to-Brachium Pulse Wave Velocity for Cardiovascular Disease Risk

**DOI:** 10.1101/2024.02.28.24303520

**Authors:** Jun Sugawara, Hirofumi Tanaka, Akira Yamashina, Hirofumi Tomiyama

## Abstract

**BACKGROUND:** Heart-brachium pulse wave velocity (hbPWV) is a measure of proximal aortic stiffness and does not require the technical placement of transducers. To characterize age-associated changes and the clinical utilities of heart-brachium pulse wave velocity (hbPWV), we conducted both cross-sectional and longitudinal analyses and used brachial-ankle pulse wave velocity (baPWV) as a comparative measure.

**METHODS:** Various measures of CVD risk factors and arterial stiffness were obtained in 7,868 adults in the cross-sectional study. In the longitudinal study, 3,710 adults were followed for 9.1 ± 2.0 years. Repeated-measures correlation (rmcorr) and regression analyses were used to characterize associations of PWVs with age and Framingham’s general CVD risk score (FRS).

**RESULTS:** In the cross-sectional study, hbPWV showed a stronger linear (r=0.796) association with age than baPWV (r=0.554). In the longitudinal study, hbPWV also showed a significantly higher rmcorr coefficient with age than baPWV (r_rm_=0.511 vs. 0.307, P<0.0001). Age-related increases in hbPWV were fairly steady starting from young adults while baPWV displayed accelerated increases with age. A receiver operating characteristic curve analysis indicated that hbPWV depicted a more robust ability to stratify general cardiovascular disease (CVD) risk compared with baPWV (AUC=0.913 vs. 0.833, P<0.0001). The results of the 10-year follow-up study were consistent with the findings of the cross-sectional investigation.

**CONCLUSIONS:** hbPWV exhibits steady and more pronounced increases from an early age and may be more effective in stratifying general CVD risk.

**NOVELTY AND RELEVANCE:** **What Is New?**

Using the cross-sectional and longitudinal study designs in a large cohort of middle-aged Japanese workers, we confirmed that hbPWV, a promising method for assessing proximal aortic stiffness, undergoes a linear augmentation with age, commencing from an early life stage onward, rendering it a potential marker for discerning CVD risk.

**What Is Relevant?**

The stiffening of the arterial wall with advancing age is particularly pronounced in the proximal aorta and contributes importantly to elevated risks of CVD, cardiovascular events, and all-cause mortalities.

**Clinical/Pathophysiological Implications?**

Early-life assessment of proximal aortic stiffening through hbPWV has the potential to mitigate the pervasive impact of CVD on a global scale.

## INTRODUCTION

The arterial system has a pivotal role called the “Windkessel” function. The arterial wall expands and recoils repeatedly with cardiac ejection from the left ventricle (LV), which minimizes energy wastage and maintains that extra component of hydraulic load as low as possible ^1–3^. Because of the prominent elastic and viscous properties and the anatomical location directly connected to the LV, the proximal aorta is considered the most important segment for the arterial buffering function. The stiffening of the arterial wall with advancing age is particularly pronounced in the proximal aorta ^4–6^ and contributes importantly to elevated risks of cardiovascular diseases (CVD), cardiovascular events, and all-cause mortalities ^7–10^. Despite recognizing that the assessment of proximal aortic stiffness would be valuable, few methodologies (e.g., magnetic resonance imaging [MRI]) are targeted for evaluations in the proximal aorta ^5,11^.

Pulse wave velocity has become the reference standard modality for assessing arterial stiffness due to a compelling association between elevated carotid-femoral pulse wave velocity (cfPWV) and risks for the development of cardiovascular disease (CVD) ^12–15^. Additionally, brachial-ankle pulse wave velocity (baPWV) is also associated with an increased risk of total cardiovascular events and all-cause mortality ^13,15,16^ and has garnered increasing acceptance ^17,18^. These measures of arterial stiffness, baPWV and cfPWV, are significantly associated with each other as they reflect overlapping regional arterial properties and functions ^19–21^. However, neither PWV measure covers the segment of the proximal aorta. In this context, heart-to-brachium pulse wave velocity (hbPWV) is a promising method ^6,22^. It does not require the technical placement of transducers. Pulse transit time can be evaluated easily by simultaneous recordings of the heart sound or ECG and brachial arterial pulse waves recorded with the high-fidelity sensor embedded in the blood pressure cuff. As an initial step to evaluate this technique, we measured the arterial path length from the heart and the brachium using MRI and constructed formulas for estimating arterial path length ^22^. A subsequent cross-sectional study employing the arterial path length estimation showed that hbPWV displayed one of the largest age-related increases compared with other measures of arterial stiffness, including cfPWV and baPWV ^6^. These results were confirmed by a 10-year follow-up investigation ^6^. However, due to the small sample size and the lack of clinical evidence, a larger-scale study focusing on clinical evidence is needed for hbPWV to gain more acceptance.

In the present study, we utilized both cross-sectional and longitudinal study designs to characterize age-related changes in hbPWV in a large cohort of middle-aged Japanese workers ^23,24^. For comparison purposes, baPWV was also assessed since this measure of arterial stiffness has been widely studied and implemented, particularly in Japanese populations. Additionally, clinical utilities of hbPWV were evaluated against general CVD risks evaluated with Framingham’s risk factors ^25^.

## METHODS

### Design and Participants

The present study was initiated on the employees working at the headquarters of a single large Japanese construction company in 2000 when the Occupational Health and Safety Law in Japan mandated annual health checkups for large companies. All company employees had to undergo annual health checkups. Participants underwent a physical examination, phlebotomy, anthropometry, blood pressure, and arterial stiffness (via PWV) measurements. Individuals with risk factors for cardiovascular disease were advised to visit the health care center within the company. They received advice for therapeutic lifestyle modifications by health professionals following the Japanese guidelines ^26–29^. Individuals requiring medications had appropriate drugs prescribed at the health care center or other clinics. The record of hbPWV measurement was not available till 2003 because of technical reasons. The available data from 2003 to 2013 were analyzed for the present study. Inclusion criteria were <75 years of age, ankle-brachial index ≥0.9, and no history of CVD. Informed consent was obtained from the study participants before participating in this study. The study was conducted with the approval of the Ethics Guidelines Committee of Tokyo Medical University (No. SH3450). Some data from this study have been published elsewhere to examine the longitudinal association of arterial stiffness (baPWV) with cardiovascular risk status ^23,24^.

### Risk Factors for Cardiovascular Disease

All the blood samples were obtained in the morning after overnight fasting. Plasma glucose concentration and serum concentrations of triglyceride, total cholesterol, and HDL-cholesterol were measured with standard enzymatic methods (Falco Biosystems Company, Ltd., Tokyo, Japan) (20). Diabetes was defined as ≥126 mg/dL of fasting glucose and/or use of insulin or oral hypoglycemic medications. Antihypertensive medication use was based on self-report. Cigarette smoking status was ascertained by self-report. Framingham’s general CVD risk score (FRS) was evaluated by sex-specific multivariable risk functions (“general CVD” algorithms) incorporated age, total and high-density lipoprotein cholesterol, systolic blood pressure, treatment for hypertension, smoking, and diabetes status ^25^.

### Cardiovascular measurements

Heart rate, BP, and PWV were measured noninvasively with the automated cardiovascular screening device (Form/ABI, Colin Company, Ltd., Komaki, Japan) equipped with an electrocardiogram, phonocardiogram, and four-extremity BP cuffs involving air-plethysmograph ^20,21,23^ after at least 5 min in the supine position in an air-conditioned room (maintained at 24°C) allocated exclusively for this study. Electrocardiographic electrodes were placed on both wrists, and a microphone for the phonocardiogram was placed on the left chest. Bilateral brachial and posterior-tibial arterial pressure waveforms were recorded for 10 seconds by occlusion cuffs connected to air-plethysmographic sensors wrapped on both arms and ankles. Pulse transit times were determined from the time delay from the proximal (e.g., brachial artery) to the distal (e.g., posterior-tibial artery) “foot” waveforms (T_ba_) for baPWV and the second heart sound to the dicrotic notch on the arterial waveforms at the right brachium (T_hb_) for hbPWV ^6,22^. Arterial path lengths for baPWV (Lba) and hbPWV (Lhb_eq1_ and Lhb_eq2_) were calculated from the following equations ^20,22^:

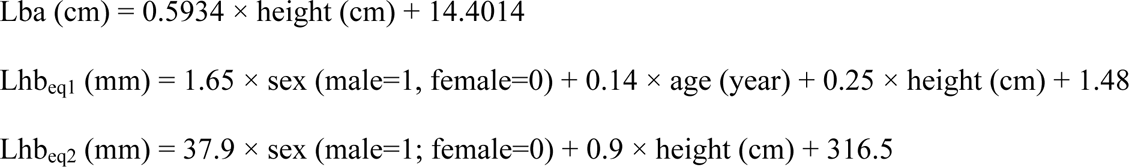

hbPWV obtained from Lhb_eq1_ and Lhb_eq2_ were reported as hbPWV_eq1_ and hbPWV_eq2_, respectively. The higher value of either the right or left sides was used for representative baPWV. Ankle-brachial index (ABI = ankle SBP/brachial SBP) was evaluated for screening for possible peripheral artery disease ^30^. Before this study, we confirmed the favorable day-to-day reproducibility of hbPWV (coefficient of variation = 4.1%) equivalent to those of heart rate, blood pressure, and baPWV (3.2, 4.7, and 3.4 %, respectively).

### Statistical Analyses

In the cross-sectional study, only data from the first visit were used for correlation analysis to avoid double counting the same participants. In the longitudinal study, data of participants who attended ≥4 examinations for ≥5 years were analyzed. The age-related changes in PWV were characterized by linear regression analyses and the repeated-measures correlation (rmcorr) using the R package ^31^. Two correlation coefficients were compared by Fisher’s z-transformation. The slope of the regression line between age and PWVs was compared among groups categorized by the initial age of participation (< 35, 35-39, 40-44, 45-49, 50-54, and ≥ 55 years) as well as PWVs by the repeated-measures ANOVA. Data obtained at the first (oldest) and final (latest) examinations were selected for determining associations between PWVs and FRS in the longitudinal study. A receiver operating characteristic (ROC) curve analysis was performed to confirm whether hbPWV stratifies a more than moderate (>10 % of 10-year risk) general CVD risk evaluated by “general CVD algorithms” ^25^ in the cross-sectional and longitudinal study designs. Cutoff points in the ROC curve were defined by the Youden index. The diagnostic accuracy of PWVs was evaluated by comparing AUC via DeLong’s test ^32^. All the analyses were conducted with RStudio (2023.09.1 Build 494, Posit Software, PBC) and Statistica (ver. 13.5.0.17, TIBCO Software Inc.). The P-values <0.05 were considered to denote statistical significance.

## RESULTS

A flowchart of the selection process for the participants is depicted in Supplemental Figure 1. A total of 8,035 employees took annual health checkups at least once throughout the observation period. One participant aged ≥75 years, 123 participants with CVD history, 42 who had ABI <0.9, and 1 with CVD history and ABI <0.9 at the initial examination were excluded. Thus, a total of 7,868 participants were analyzed for the cross-sectional analysis (Table 1). Among them, 3,710 participants (46.2%) met the inclusion criteria for the longitudinal analysis (attended ≥4 examinations for ≥5 years). Three hundred seventy-eight participants did not have FRS data due primarily to no phlebotomy. The average ages of the participants at the initial (oldest) and final (latest) tests were 40.5±8.6 and 48.5±8.6 years. The average duration and number of health checkups were 9.1 ± 2.0 years and 6.7 ± 1.7 times, respectively.

**Table 1.** Subjects’ characteristics in the cross-sectional and longitudinal studies.

| Variables | Cross-sectional study | Longitudinal study |  |
| --- | --- | --- | --- |
|  |  | Initial examination | Final examination |
| N, men/women | 6,683 / 1,185 | 3,110/600 | 3,110/600 |
| Age, years | 42.3 ± 10.3 | 40.5 ± 8.6 | 48.5 ± 8.6 |
| Height, cm | 169.2 ± 7.5 | 169.5 ± 7.4 | 169.1 ± 7.5 |
| Weight, kg | 67.7 ± 12.0 | 67.8 ± 11.9 | 68.4 ± 12.1 |
| BMI, kg/m<sup>2</sup> | 23.6 ± 3.3 | 23.5 ± 3.3 | 23.8 ± 3.3 |
| TC, mg/dl | 200.7 ± 34.7 | 199.8 ± 34.8 | 199.8 ± 34.8 |
| HDL-C, mg/dl | 59.3 ± 14.8 | 59.3 ± 14.7 | 59.3 ± 14.7 |
| Triglyceride, mg/dl | 121.5 ± 101.2 | 118.2 ± 95.2 | 118.2 ± 95.2 |
| Blood glucose, mg/dl | 91.8 ± 16.9 | 90.8 ± 12.9 | 90.8 ± 12.9 |
| HbA1c, % | 5.1 ± 0.6 | 5.0 ± 0.5 | 5.05 ± 0.54 |
| FRS, points | 7.4 ± 6.5 | 6.5 ± 5.9 | 9.4 ± 5.8 |
| Heart rate, beat/min | 65.9 ± 10.1 | 65 ± 10 | 65 ± 9 |
| Systolic BP, mmHg | 127.4 ± 16.9 | 126 ± 15 | 123 ± 15 |
| Diastolic BP, mmHg | 77.3 ± 11.9 | 76 ± 11 | 77 ± 11 |
| baPWV, cm/sec | 1308 ± 223 | 1269 ± 181 | 1343 ± 222 |
| hbPWV, cm/sec | 740 ± 156 | 715 ± 134 | 806 ± 145 |
| BP treatment, n (%) | 949 (12.1) | 154 (4.1) | 519 (14.0) |
| DM treatment, n (%) | 271 (3.4) | 37 (1.0) | 141 (3.8) |
| HL treatment, n (%) | 451 (5.7) | 49 (1.3) | 236 (6.4) |
| Smoking habit, n (%) | 3,894 (49.5) | 1806 (48.7) | 1806 (48.7) |
| Observation period, years | 4.5 ± 4.0 | 9.1 ± 2.0 | |
| Data count, n | 4.1 ± 2.9 | 6.7 ± 1.7 | |

Data are mean and 95% confidential interval. BMI: body mass index; TC: total cholesterol; HDL-C: high-density lipoprotein cholesterol; HbA1C: hemoglobin A1C; FRS: Framingham risk score; BP: blood pressure; baPWV: brachial-ankle pulse wave velocity; hbPWV: heart-brachium pulse wave velocity; DM: diabetes mellitus; HL: hyperlipidemia.

### Aging-related increases in PWV

In the cross-sectional observation, hbPWV_eq1_ showed stronger linear associations with age in men (r = 0.796, P<0.0001), women (r = 0.748, P<0.0001), and whole participants (r = 0.796, P<0.0001) (Figure 1A). hbPWV_eq2_ also showed stronger linear associations with age in men (r = 0.747, P<0.0001), women (r = 0.681, P<0.0001), and whole participants (r = 0.746, P<0.0001) (Figure 1B). These correlation coefficients were significantly larger than those of baPWV (men: r = 0.523, P<0.0001; women: r = 0.638, P<0.0001; whole participants: r = 0.554, P<0.0001) (Figure 1C).

**Figure 1.**
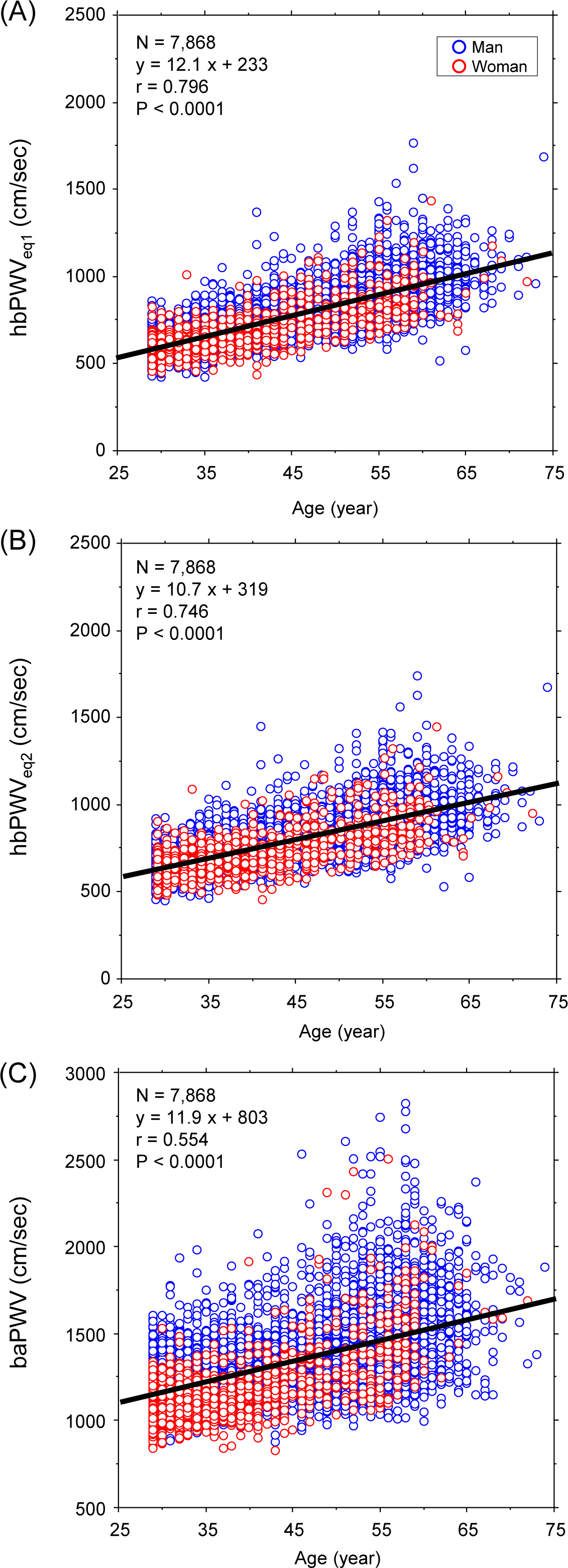
Simple correlation analyses between age and pulse wave velocity (PWV) indices in the cross-sectional study. Blue and red open circles indicate man and women, respectively. A black line is the linear regression line of whole subjects.

In the longitudinal observation, hbPWV_eq1_ and hbPWV_eq2_ showed significant rmcorr coefficients with age in men (r_rm_ = 0.516 and 448, P<0.0001 for both), women (r_rm_ = 0.485 and and 396, P<0.0001 for both), and whole participants (r_rm_ = 0.511 and 439, P<0.0001 for both) (Figure 2A and B). These r_rm_ were significantly than those of baPWV (men: r_rm_ = 0.308, P<0.0001; women: r_rm_ = 0.300, P<0.0001; whole participants: r_rm_ = 0.306, P<0.0001) (Figure 2C).

**Figure 2.**
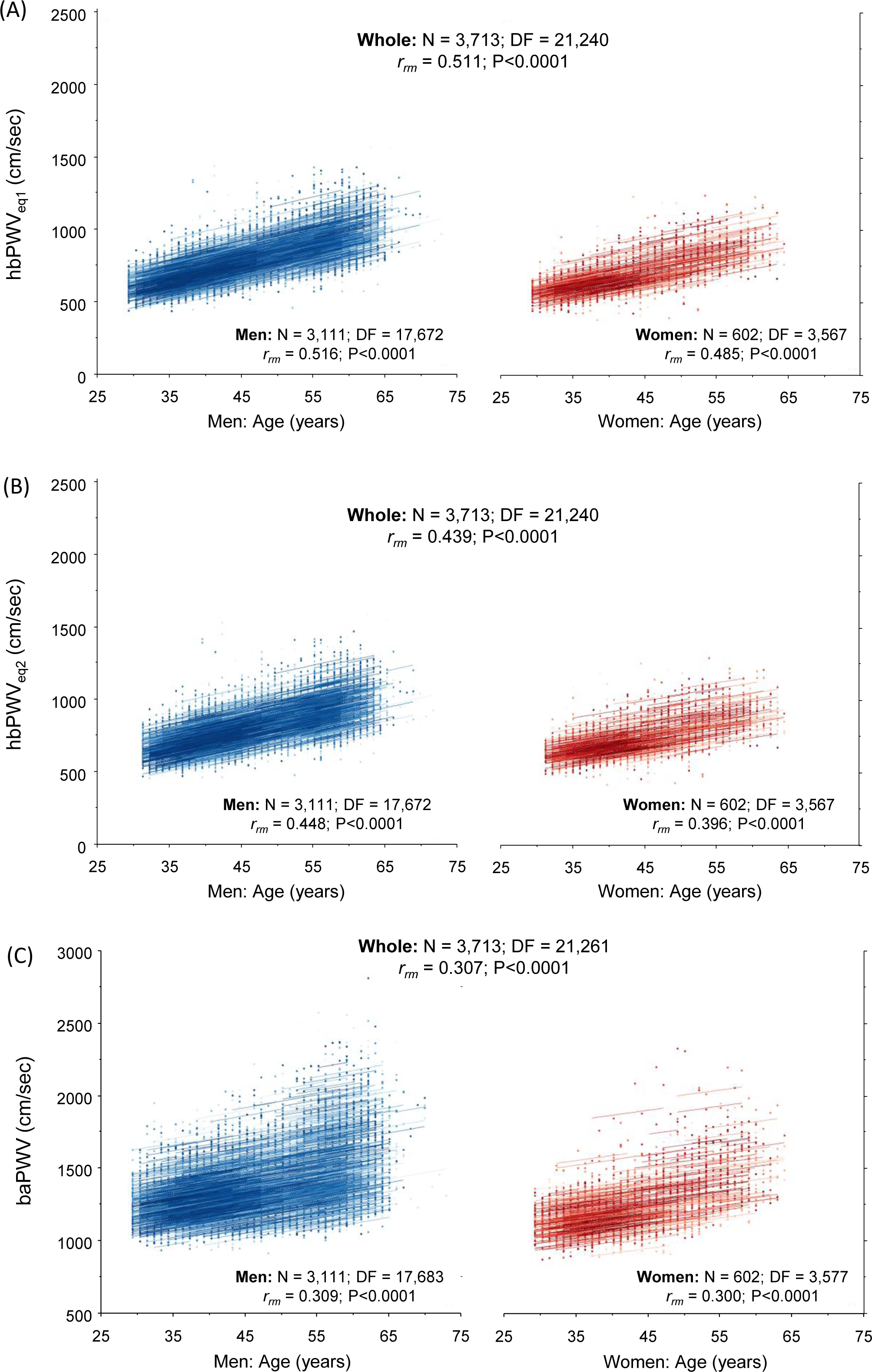
Repeated measures correlation plots between age and pulse wave velocity (PWV) indices. Lines represent intra-individual linear regression lines for each participant. DF, degrees of freedom; rrm, the coefficient of repeated measures correlation.

Figure 3 depicts comparisons of the slope of the regression line between age and PWVs among groups categorized by the initial age of participation. The age-related slope of baPWV gradually increased with the initial age, whereas that of hbPWVs were fairly steady starting from young adults. the linear regression slopes of hbPWVs were significantly larger in the age groups of <40 years but became significantly smaller in the age groups of ≥50 years than those of baPWV.

**Figure 3.**
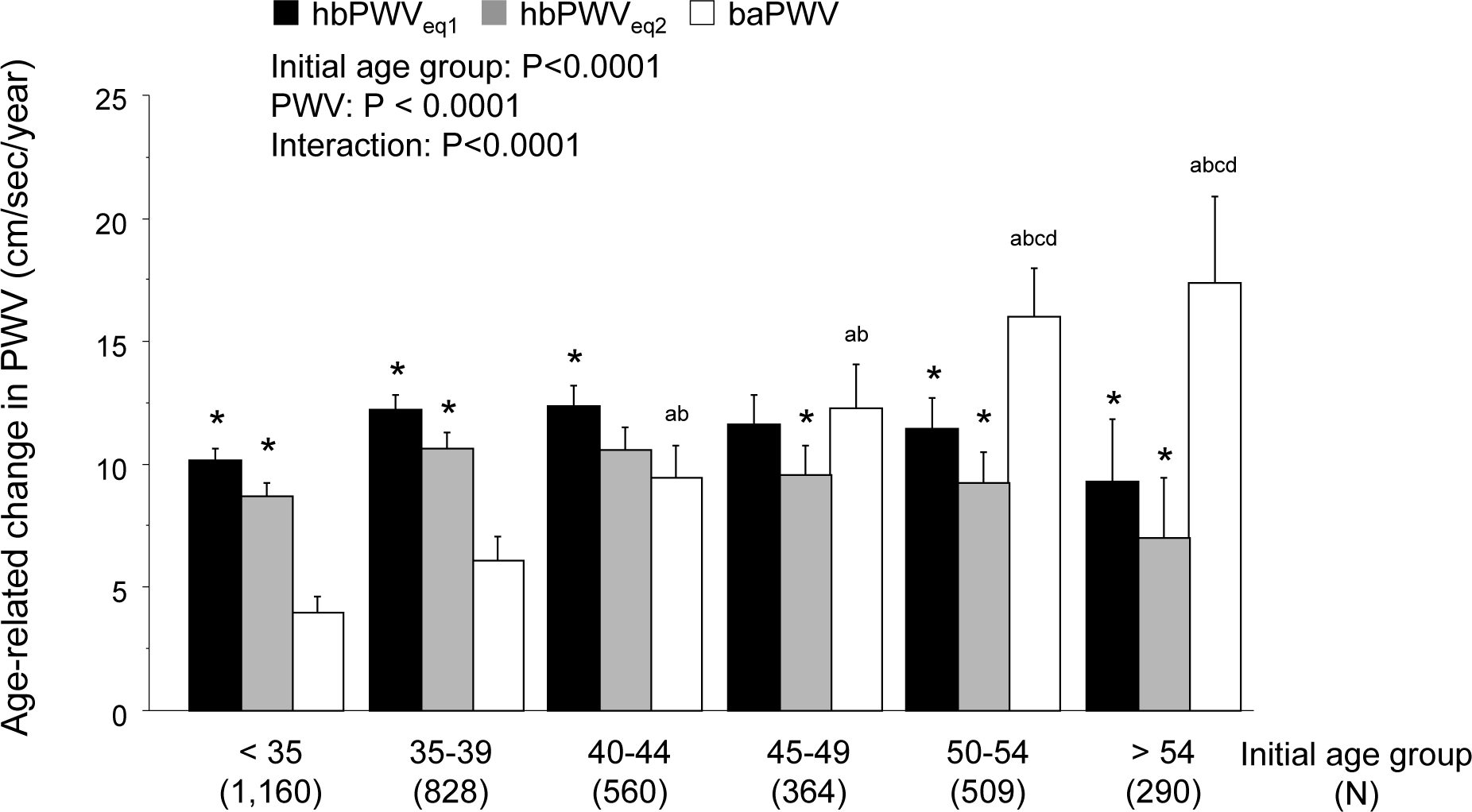
The age-related regression slopes of pulse wave velocity (PWV) indices. a) P<0.0001 vs. <35 years of the initial age group. b) P<0.0001 vs. 35-39 years of the initial age group. c) P<0.0001 vs. 40-44 years of the initial age group. d) P<0.0001 vs. 45-49 years of the initial age group. *P<0.0001 vs. baPWV. Error bars indicate 95% confidential interval.

### Relation to CVD risk

In the cross-sectional study, 2,399 participants were classified into the more than moderate general CDV risk group. hbPWV_eq1_, hbPWV_eq2_, and baPWV correlated with FRS logarithmically in men (r = 0.755, 0.720, and 0.601, respectively), women (r = 0.734, 0.684, and 0.746, respectively), and whole participants (r = 0.755, 0.718, and 0.659, respectively) (all P < 0.0001) (Figure 4A-C). As shown in Figure 4D, baPWV stratified high CVD risk with 0.833 of the AUC. Compared with baPWV, hbPWV_eq1_ and hbPWV_eq2_ exhibited more robust ability to stratify general CVD risk (AUC = 0.913 and 0.896, P<0.0001 for both via DeLong’s test, Figure 4D).

**Figure 4.**
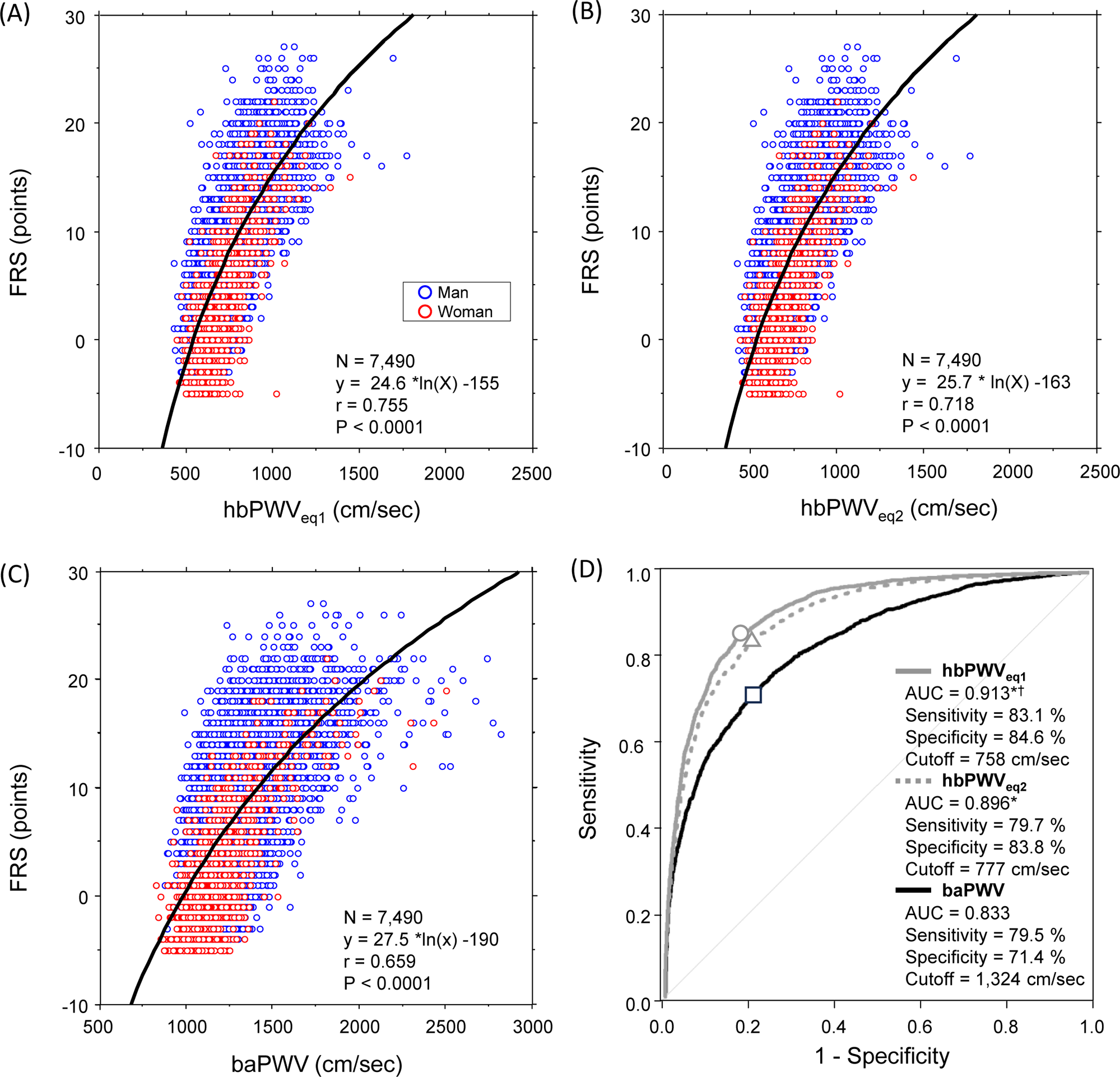
Logarithmic correlation plots between pulse wave velocity (PWV) indices and Framingham risk score (FRS) in the cross-sectional study (Panel A-C). Panel D indicates the results of a receiver operating characteristic (ROC) curve analysis. * Significant difference in AUC of baPWV. ^†^ Significant difference in AUC of hbPWV_eq2_. Open circle, triangle, and square indicate cutoff point of hbPWV_eq1_, hbPWV_eq2_, and baPWV, respectively.

Figure 5 displays results of correlation and ROC curve analyses between PWVs and FRS at the initial and final examinations in the longitudinal observation. The more than moderate-risk holders were 1,124 at the initial examination and 1,740 at the last. In pooled participants, stronger logarithmic correlations of PWVs with FRS were seen at both the initial and final examinations (hbPWV_eq1_: r = 0.708 and 0.698; hbPWV_eq2_: r = 0.667 and 0.662; baPWV: r = 0.600 and 0.658). The plot and regression lines almost overlapped in each PWV at the initial and final examination. Scatter plots are displayed at the initial and final examination separately in supplemental Figure 2. AUC of hbPWVs were significantly decreased at the final examination compared with those at the initial (results), whereas AUC of baPWV was significantly increased. hbPWV_eq1_ and hbPWV_eq2_ exhibited significantly higher AUC compared with baPWV (0.891 and 0.872 vs. 0.788, P<0.0001 for both) at the initial examination, whereas no differences in AUC between hbPWVs and baPWV were observed at the final examination.

**Figure 5.**
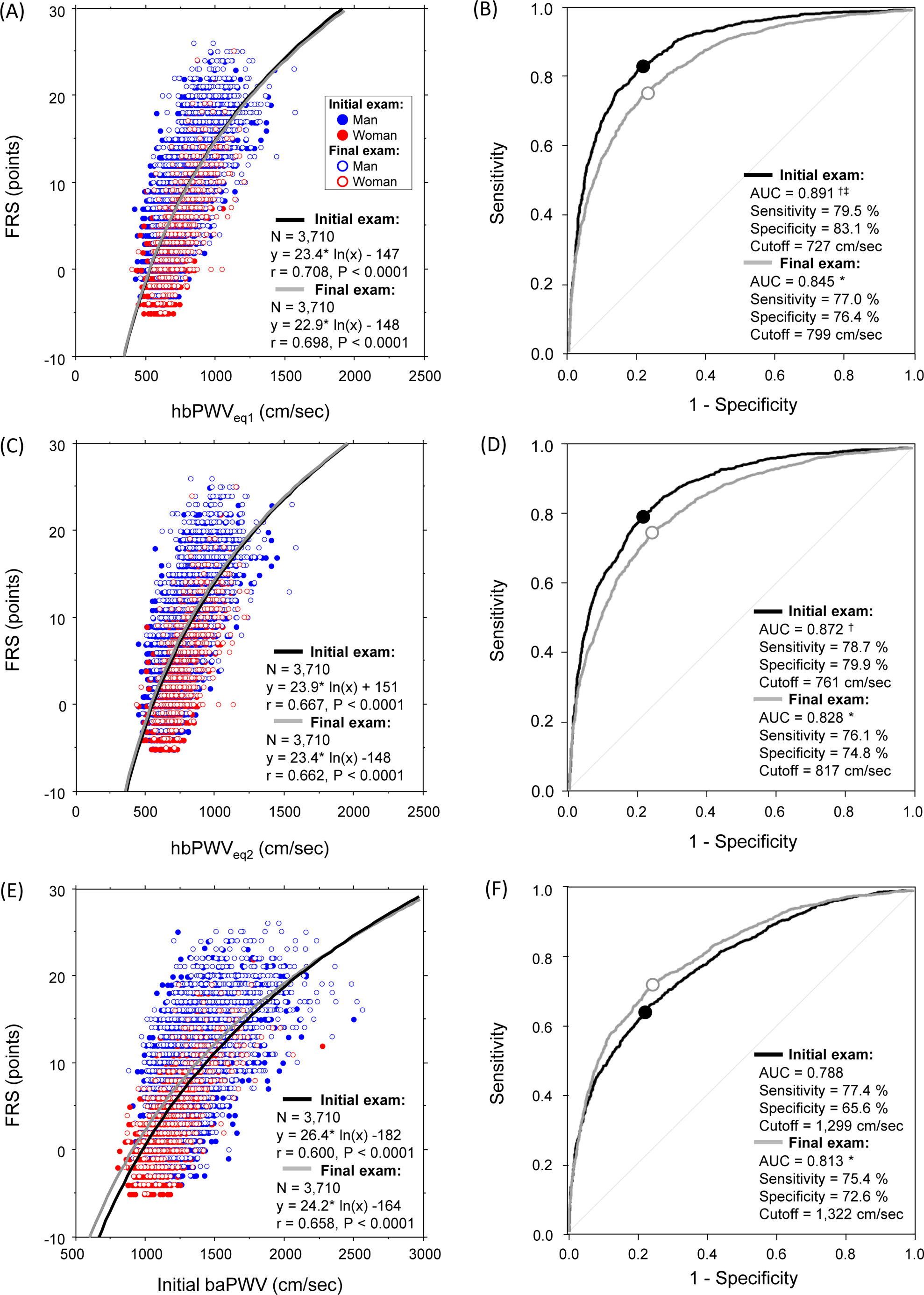
Logarithmic correlation plots between pulse wave velocity (PWV) indices and Framingham risk score (FRS) in the longitudinal study (left panels) and the results of a receiver operating characteristic (ROC) curve analysis (right panels). Open and filled circles indicate cutoff point of at the initial and final examination, respectively. *Significant difference vs. the initial examination. †Significant difference vs. baPWV. ‡Significant difference vs. hbPWVeq2.

## DISCUSSION

The salient findings of the present study are as follows. First, compared with baPWV, hbPWV exhibited stronger associations with age and FRS. Second, unlike baPW that displayed gradually accelerated increases with advancing age, hbPWV demonstrated steady and consistent age-related increases throughout the lifespan. Third, the ROC curve analyses indicated that hbPWV depicted a more robust ability to stratify general cardiovascular disease (CVD) risk compared with baPWV. The results of the 10-year follow-up study were consistent with the findings of the cross-sectional investigation, irrespective of the level of risk and the temporal dimension (e.g., the initial and final examinations). We conclude that hbPWV should be considered a good marker for discerning CVD risk.

Heart-to-brachium pulse wave velocity (hbPWV) has been proposed as an indicator of proximal aortic stiffness ^6,22^. However, its adoption as a vascular biomarker has been hindered by the uncertainty in assessing arterial path length ^33^. As the initial step to better define hbPWV, we proposed and validated a simple equation developed through the application of MRI ^22^. The subsequent study showed that hbPWV recalculated using the newly-created equation displayed a pronounced age-associated increase and significant associations with central hemodynamic factors associated with CVD risk ^6^. The present cross-sectional and longitudinal study presents the data that further supports and substantiates the clinical relevance of hbPWV.

### Aging-related increases in PWVs

It is widely acknowledged that arterial stiffness increases with the progression of age ^19–21,34^. The majority of the currently available evidence stems from cross-sectional observations and does not capture intra-individual changes in arterial stiffness with aging. The present study is an attempt to close the gap in the field. When conventional correlation analysis is applied to non-independent observations such as repeated-measures aggregated data, biased results might emerge due to the violation of independence and/or differing patterns between subjects versus within subjects ^31^. Therefore, we employed the repeated measures correlation (rmcorr), a statistical technique for determining the common within-individual association for paired measures assessed on more occasions for multiple individuals ^31^. The rmcorr coefficients with age were substantially higher for hbPWV than for baPWV, irrespective of the arterial length equations with or without age. Additionally, we observed a relatively smaller variance of the aging slope for hbPWV across the entire lifespan. The longitudinal observations were consistent in finding the steady and consistent age-related increases in hbPWV throughout the lifespan.

The age-related slope of baPWV exhibited attenuation in younger generations highlighting more pronounced increases after reaching middle age. This characteristic can be ascribed to the methodology employed in baPWV measurement. The arterial path length covered in baPWV encompasses both central elastic arteries (e.g., abdominal aorta) and peripheral muscular arteries (e.g., leg arteries). The stiffness of muscular arteries is generally greater than that of central elastic arteries in young and middle-aged adults and does not change much with aging ^35,36^. In contrast, central elastic arteries progressively stiffen from an early age ultimately surpassing the stiffness levels of muscular arteries around 50 years ^6,34^. This dynamic interplay engenders impedance matching between central arteries (such as the abdominal aorta) and peripheral conduit arteries (such as the iliac and femoral arteries) ^37^. Such impedance matching, in turn, might mitigate wave reflection and promote the smooth forward transition of pulse waves in phase ^3^. Consequently, this intricate mechanism is likely instrumental in the gradual escalation of baPWV with aging observed in older populations. Although the arterial trajectory of hbPWV also includes both elastic (i.e., the ascending aorta) and muscular (i.e., the subclavian, axial, and brachial arteries) arteries, the proportion of elastic arteries in this arterial trajectory is much greater. In the present study, hbPWV exhibited a sustained elevation even during the early stages of life. A previous study using MRI indicated a substantial decrease in distensibility of the ascending aorta even in younger age ^5^. It is likely that the age-related alterations in hbPWV may mirror the age-induced stiffening of the proximal aorta.

### Relation to CVD risk

The elevated arterial stiffness is linked to cardiovascular events and all-cause mortality ^12–16^. Consequently, integrating assessments of arterial stiffness into routine clinical practice holds promise for enhancing and optimizing CVD risk management ^38^. We ^39^ and others ^40^ substantiated that baPWV can act as a discriminator for identifying individuals at elevated risk of CVD. In the present study, we identified the steady and consistent age-related increases in hbPWV from the early life stage, implying the potential of a more sensitive marker for early detection of CVD risk. Accordingly, we determined whether hbPWV could stratify moderate CVD risk (>10% of 10-year risk). hbPWV consistently demonstrated a superior AUC compared with baPWV, irrespective of the temporal dimension (e.g., the initial and final examinations). Furthermore, AUC of hbPWV was higher at the initial than the final testing periods. These findings collectively indicate hbPWV as a more effective screening tool for ascertaining general CVD risk, especially in the early life stage.

While the precise mechanistic underpinnings remain elusive, there are a number of supporting evidence. First, hbPWV mainly reflects proximal aortic stiffness, notably the most elastic segment directly linked to the LV. Its role in attenuating cyclic mechanical forces generated by cardiac pulsations is particularly recognized. This critical segment is omitted in baPWV. Second, unlike baPWV, that is measured at the end of diastole via the foot-to-foot method, the pulse transition time for hbPWV is measured in systole and aligns with aortic pressure and LV afterload. Indeed, our previous study ^6^ revealed that the longitudinal changes in hbPWV exhibited a more robust correlation with corresponding alterations in aortic hemodynamic variables (e.g., aortic systolic blood pressure, augmentation pressure, and augmentation index), which are independent predictors of future CVD events and all-cause mortality ^12,41,42^.

### Study Limitations

As in any research studies, there are many limitations associated with the present study. First, following the health checkups, participants were advised to modify their lifestyle habits, including regular physical activity or dietary intake, during the observation. Given that we have no information or control over these individual choices, the potential impact of these variables on the observed alterations in PWVs remains unknown. Second, because one of the equations to estimate arterial path length for hbPWV involved age, its strong correlation with age is not surprising. However, the results were not different from hbPWV calculated using the arterial length equation that did not involve age. Third, the notably smaller representation of women compared with men in our study precluded a comprehensive examination of individual PWV changes with aging from a sex-specific standpoint. Last, the associations between hbPWV and clinical events or endpoints are unknown. The accuracy of predicting future events necessitates validation through prospective studies.

## CONCLUSION

To further evaluate the promising modality of hbPWV in evaluating central elastic artery stiffness, we conducted combined cross-sectional and longitudinal analyses to determine age-associated changes and general CVD risk. Compared with baPWV, hbPWV demonstrated more robust associations with age and FRS. Particularly noteworthy was the feature of hbPWV to display significantly greater age-related increases even in young adults. In contrast, baPWV exhibited a gradually accelerated increment with advancing age. The ROC curve analysis further substantiated superior efficacy of hbPWV in stratifying general CVD risk compared with baPWV. Taken together, our present findings suggest that hbPWV undergoes a linear augmentation with age, commencing from an early life stage onward, rendering it a potential marker for discerning CVD risk.

## PERSPECTIVES

Our results do not support the notion that hbPWV is a surrogate marker for baPWV even though both methods are easy to implement in routine clinical settings as they do not require technical placement of transducers. The segments of arterial paths covered by baPWV and hbPWV are very different. Consequently, these measures may offer distinct insights into various arterial functions. baPWV has gained recognition as a convenient screening tool for hypertension-mediated organ damage ^17,18^. hbPWV may reflect age-related changes in the proximal aortic properties, such as Windkessel function, starting from the early life stage. The Network for Research in Vascular Ageing recommended integrating assessment of sub-clinical arteriosclerosis (via baPWV), advanced atherosclerosis (via ABI and systolic interarm blood pressure difference), and central hemodynamics associated with LV afterload (via pulse wave analysis) to gain a holistic snapshot of a cardiovascular health management ^38^. Therefore, incorporating hbPWV measurement into this framework enables a more comprehensive clinical assessment and management of CVD.

## Data Availability

The data that support the findings of this study are available on request from the corresponding author, [HT]. The data are not publicly available due to restrictions (e.g. their containing information that could compromise the privacy of research participants).

---

**Supplemental Figure 1.** The schematic representation of the participant selection process.

**Supplemental Figure 2.** Logarithmic correlation plots between pulse wave velocity (PWV) indices and Framingham risk score (FRS) at the initial (left panels) and final (right panels) examination.

